# AN INTERACTIVE COVID-19 MOBILITY IMPACT AND SOCIAL DISTANCING ANALYSIS PLATFORM

**DOI:** 10.1101/2020.04.29.20085472

**Authors:** Lei Zhang, Sepehr Ghader, Michael L. Pack, Chenfeng Xiong, Aref Darzi, Mofeng Yang, QianQian Sun, AliAkbar Kabiri, Songhua Hu

## Abstract

The research team has utilized privacy-protected mobile device location data, integrated with COVID-19 case data and census population data, to produce a COVID-19 impact analysis platform that can inform users about the effects of COVID-19 spread and government orders on mobility and social distancing. The platform is being updated daily, to continuously inform decision-makers about the impacts of COVID-19 on their communities using an interactive analytical tool. The research team has processed anonymized mobile device location data to identify trips and produced a set of variables including social distancing index, percentage of people staying at home, visits to work and non-work locations, out-of-town trips, and trip distance. The results are aggregated to county and state levels to protect privacy and scaled to the entire population of each county and state. The research team are making their data and findings, which are updated daily and go back to January 1, 2020, for benchmarking, available to the public in order to help public officials make informed decisions. This paper presents a summary of the platform and describes the methodology used to process data and produce the platform metrics.

## 1. INTRODUCTION

Informed decision-making requires data. In the case of COVID-19, no previous pandemic had such a big universal impact on societies in the modern history, as a results historic data lacked key information on how people react to such a universal pandemic and how the virus impacts economies and societies. Data-driven decision-making becomes a challenge in such an unprecedented event. Thanks to the technology, we now have an enormous amount of observed data collected by mobile devices amid pandemic. We can now utilize this data to learn more about the various impacts of a pandemic on our lives, make informed decisions to fight the current invisible enemy, and be better prepared the next time such pandemics happen. Our research team has utilized a national set of privacy-protected mobile device location data and produced a COVID-19 Impact Analysis Platform to provide comprehensive data and insights on COVID-19’s impact on mobility, economy, and society.

Mobile device location data are becoming popular for studying human behavior, specially mobility behavior. Earlier studies with mobile device location data were mainly using GPS technology, which is capable of recording accurate information including, location, time, speed, and possibly a measure of data quality ^1^. Later, mobile phones and smartphones gained popularity, as they could enable researchers to sudy individual-level mobility patterns ^2–4^. Other emerging mobile device location data sources such as call detail record (CDR) ^5–7^, Cellular network data ^8^, and social media location-based services ^9–13^ have also been used by the researchers to study mobility behavior. Mobile device location data has proved to be a great asset for decision-makers amid the current COVID-19 pandemic. Many companies such as Google, Apple, or Cuebiq have already utilized location data to produce valuable information about mobility and economic trends ^14–16^. Researchers have also utilized mobile device location data for studying COVID-19- related behavior ^17,18^.

Non-pharmaceutical interventions such as social distancing are important and effective tools for preventing virus spread. One of the most recent studies projected that the recurrent outbreaks might be observed this winter based on pharmaceutical estimates on COVID-19 and other coronaviruses, so prolonged or intermittent social distancing may be required until 2022 without any interventions ^19^, highlighting the importance of improving our understanding about individual’s reaction to social distancing. Researchers have highlighted the importance of social distancing in disease prevention through modeling and simulation ^20–23^. The simulation models assume a level of compliance, which can now be validated through observed data. Our current platform utilizes mobile device location data to provide observed data and evidence on social distancing behavior and the impact of COVID-19 on mobility. We used daily feeds of mobile device location data, representing movements of more than 100 Million anonymized devices, integrated with COVID-19 case data from John Hopkins University and census population data to monitor the mobility trends in United States and study social distancing behavior ^24^ In the next section we describe the methodology used to process the anonymized location data and produce the metrics that are available on the platform. The methodology section is followed by a brief overview of the platform. The last section presents concluding remarks.

## 2. METHODOLOGY

The research team first integrated and cleaned location data from multiple sources representing person and vehicle movements in order to improve the quality of our mobile device location data panel. We then clustered the location points into activity locations and identified home and work locations at the census block group (CBG) level to protect privacy. We examined both temporal and spatial features for the entire activity location list to identify home CBGs and work CBGs for workers with a fixed work location. Next, we applied previously developed and validated algorithms ^25^ to identify all trips from the cleaned data panel, including trip origin, destination, departure time, and arrival time. Additional steps were taken to impute missing trip information for each trip, such as trip purpose (e.g., work, non-work), point-of-interest visited (restaurants, shops, etc.), travel mode (air, rail, bus, driving, biking, walking, and others), trip distance (airline and actual distance), and socio-demographics of the travelers (income, age, gender, race, etc.) using advanced artificial intelligence and machine learning algorithms. If an anonymized individual in the sample did not make any trip longer than one-mile in distance, this anonymized individual was considered as staying at home. A multi-level weighting procedure expanded the sample to the entire population, using device-level and trip-level weights, so the results are representative of the entire population in a nation, state, or county. The data sources and computational algorithms have been validated based on a variety of independent datasets such as the National Household Travel Survey and American Community Survey, and peer reviewed by an external expert panel in a U. S. Department of Transportation Federal Highway Administration’s Exploratory Advanced Research Program project, titled “Data analytics and modeling methods for tracking and predicting origin-destination travel trends based on mobile device data” ^25^. Mobility metrics were then integrated with COVID-19 case data, population data, and other data sources. **Figure 1** shows a summary of the methodology.

**Figure 1.**
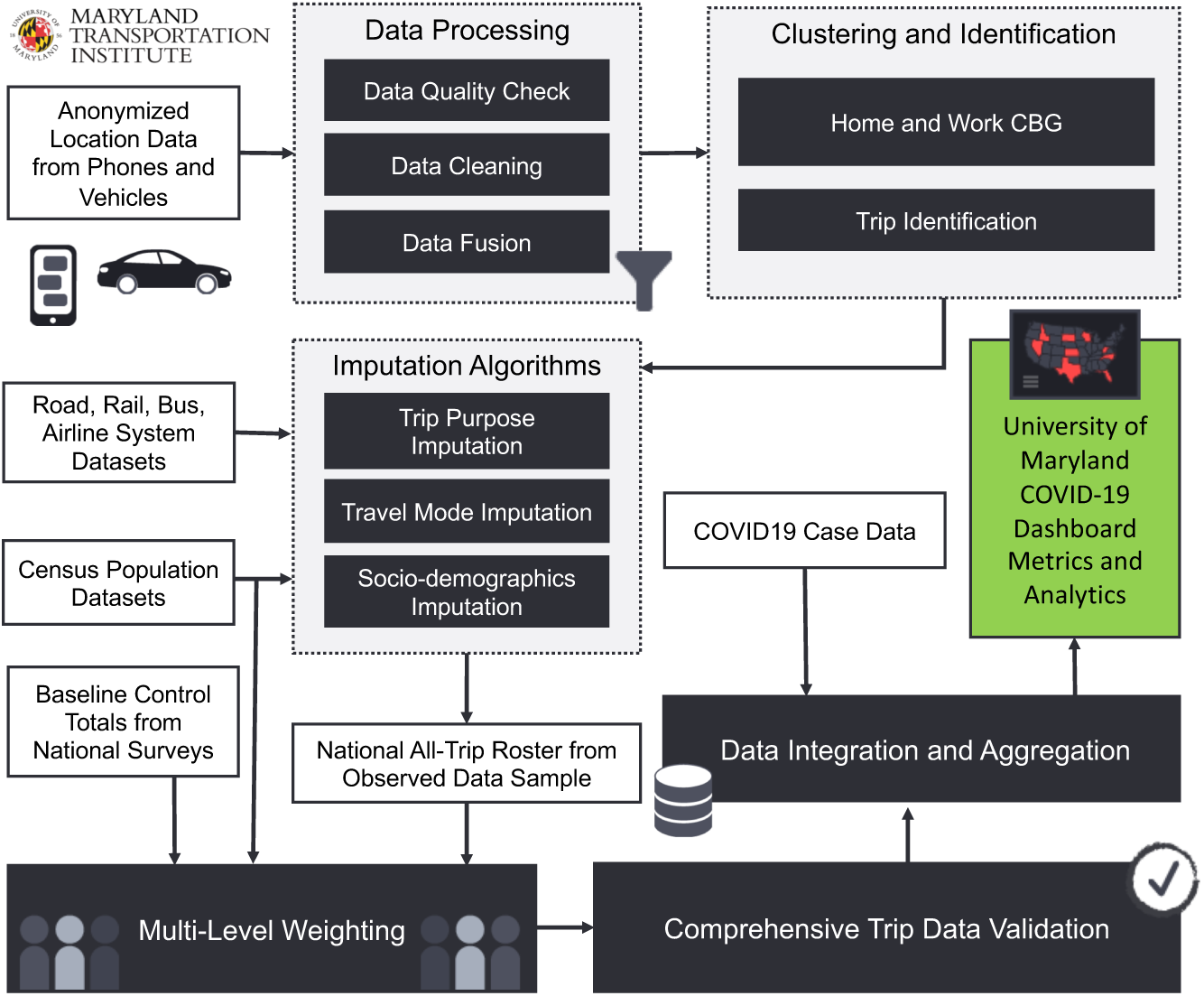
Methodology

### 2.1. Trip Identification

Trips are the unit of analysis for almost all transportation applications. Traditional data sources such as travel surveys include trip-level information. The mobile device location data, on the other hand, do not directly include trip information. Location sightings can be continuously recorded while a device moves, stops, stays static, or starts a new trip. These changes in status are not recorded in the raw data. As a result, researchers must rely on trip identification algorithms to extract trip information from the raw data. Basically, researchers must identify which locations form a trip together. The following subsections describe the steps our research team took to identify trips. The algorithm runs on the observations of each device separately.

#### 2.1.1. Pre-Processing

First, all device observations are sorted by time. The trip identification algorithm assigns a hashed ID to every trip it identifies. The location dataset may include many points that do not belong to any trips. The algorithm assigns “0” as the trip ID to these points to identify them as static points. for every observation, we compute the distance, time, and speed between the point and its previous and next points if exist.

The trip identification algorithm has three hyper-parameters: distance threshold, time threshold, and speed threshold. The speed threshold is used to identify if an observation is recorded on the move. The distance and time thresholds are used to identify trip ends. At this step, the algorithm identifies the device’s first observation with *speed from ≥ speed threshold*. This identified point is on the move, so a hashed trip ID is generated and assigned to this point. All points recorded before this point, if exist, are set to have “0” as their trip ID. Next, the recursive algorithm identifies if the next points are on the same trip and should have the same trip ID.

#### 2.1.2. Recursive Algorithm

This algorithm checks every point to identify if they belong to the same trip as their previous point. If they do, they are assigned the same trip ID. If they do not, they are either assigned a new hashed trip id (when their *speed from ≥ speed threshold)* or their trip ID is set to “0” (when their *speed from < speed threshold*). Identifying if a point belongs to the same trip as its previous point is based on the point’s “speed to”, “distance to” and “time to” attributes. If a device is seen in a point with *distance to ≥ distance threshold* but is not observed to move there *(speed to < speed threshold)*, the point does not belong to the same trip as its previous point. When the device is on the move at a point *(speed to ≥ speed threshold)*, the point belongs to the same trip as its previous point; but when the device stops, the algorithm checks the radius and dwell time to identify if the previous trip has ended. If the device stays at the stop (points should be closer than the distance threshold) for a period of time shorter than the time threshold, the points still belong to the previous trip. When the dwell time reaches above the time threshold, the trip ends, and the next points no longer belong to the same trip. The algorithm does this by updating “time from” to be measured from the first observation in the stop, not the point’s previous point. The algorithm may identify a local movement as a trip if the device moves within a stay location. To filter out such trips, all trips that are within a static cluster and all trips that are shorter than 300 meters are removed.

#### 2.1.3. Validation

**Figure 2** and **Figure 3** show the validation of this algorithm by running the algorithm on a sample of national mobile device location data and comparing the trip lengths and travel times with the reported travel distances and travel times from the 2017 national household travel survey (NHTS 2017). A satisfactory match is observed between the two datasets.

**Figure 2.**
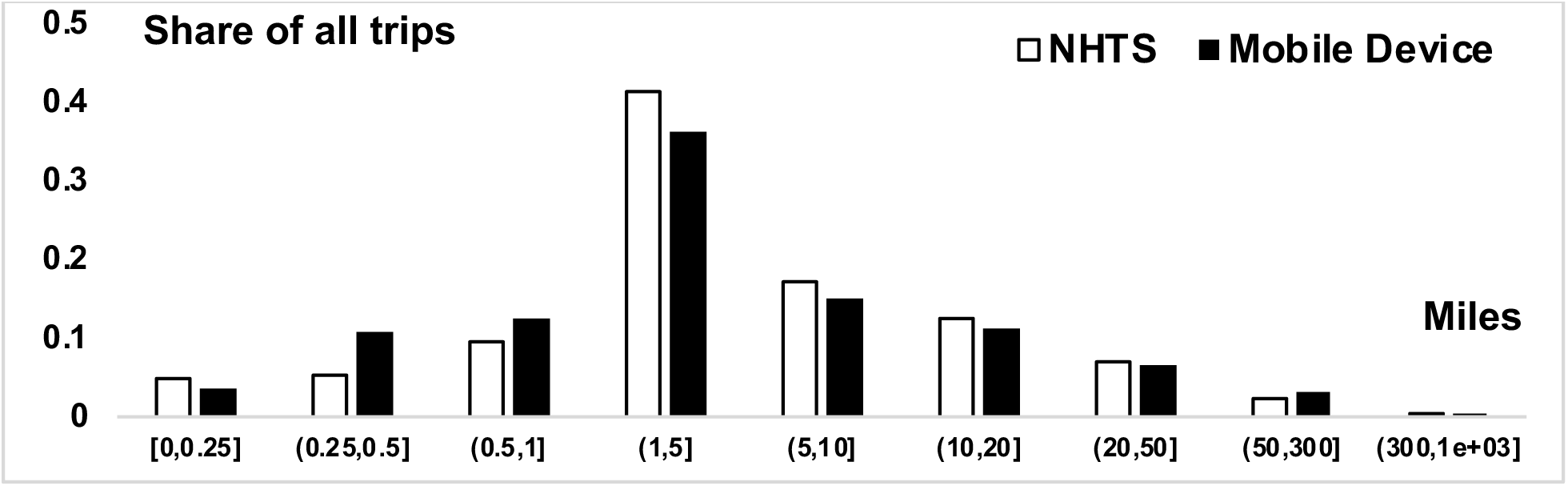
Distance validation of the trip identification algorithm against NHTS2017

**Figure 3.**
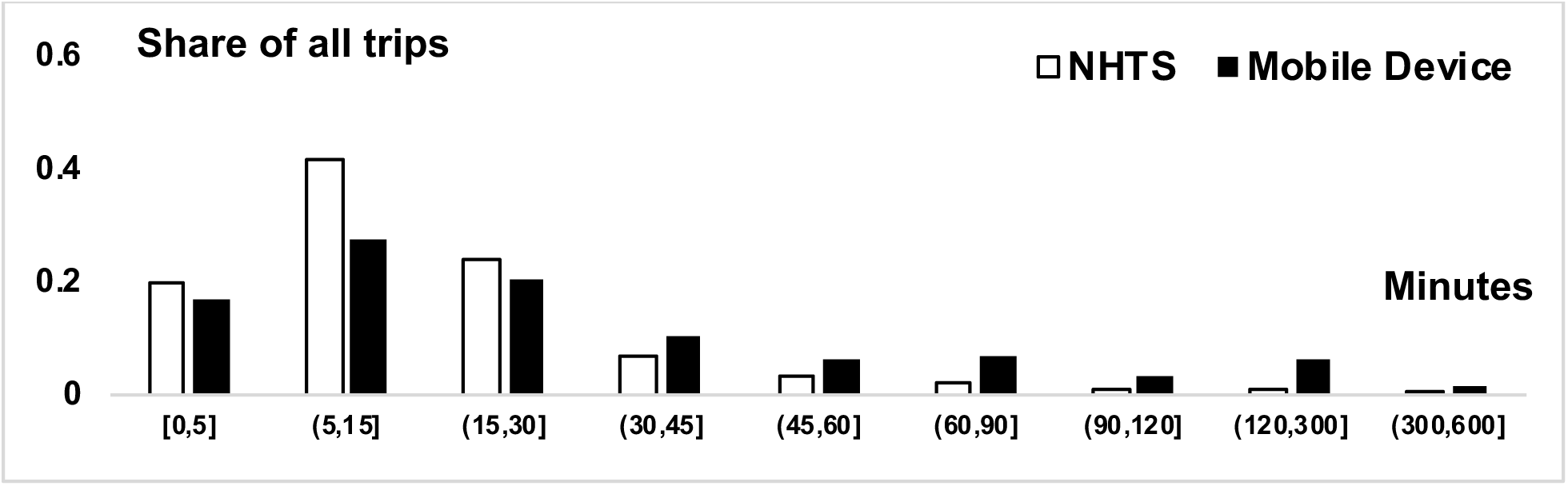
Travel time validation of the trip identification algorithm against NHTS2017

### 2.2. Activity Identification

We first identify all activity points. Then, based on the temporal and spatial distribution of activity points, we identify the home census block group (CBG) and the work CBG.

#### 2.2.1. Activity Clustering

The algorithm starts by clustering all device observations into activity locations using HDBSCAN^26^ clustering algorithm. This step takes the cleaned multi-day location data as input and applies an iterative algorithm until no cluster has a radius larger than two miles. The iterative algorithm consists of two parts: HDBSCAN based on a minimum number of point parameters and filtering non-static clusters based on time and speed checks. After finalizing the potential stay clusters, the framework combines nearby clusters to avoid splitting a single activity **(Figure 4)**.

**Figure 4.**
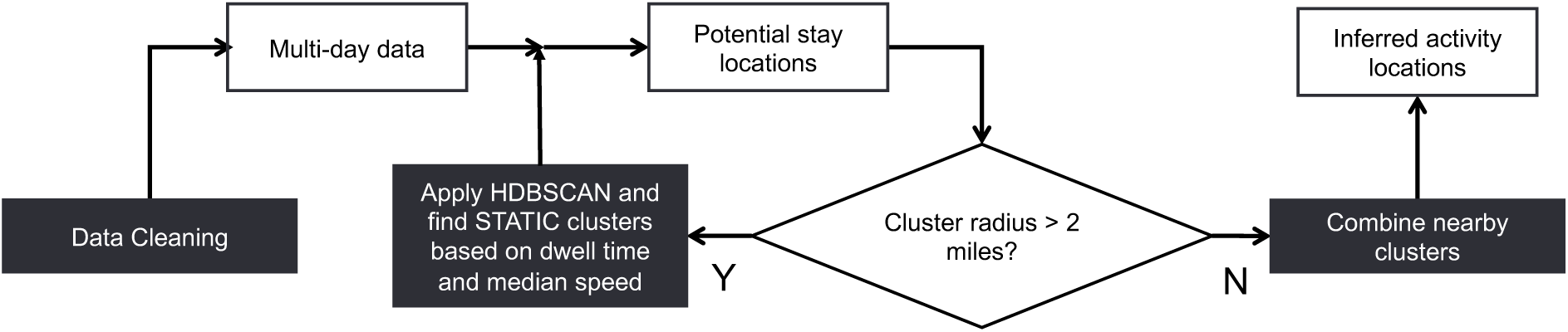
Activity clustering methodology

#### 2.2.2. Home and work CBG Identification

**Figure 5** shows the methodology for home and work CBG identification. Instead of setting a fixed time period for each type, e.g., 8pm to 8am as the study period for home CBG identification and the other half day for work CBG identification, the framework examines both temporal and spatial features for the entire activity location list. The benefits are two-fold: the results for workers with flexible or opposite work schedules would be more accurate and the employment type for each device could be detected simultaneously. **Figure 6** shows the validation of home and work location imputations, by comparing the distance from home to work between longitudinal employer-household dynamics (LEHD) data and the imputed locations for a set of mobile device location data for the Baltimore metropolitan area. We can observe a satisfactory match.

**Figure 5.**
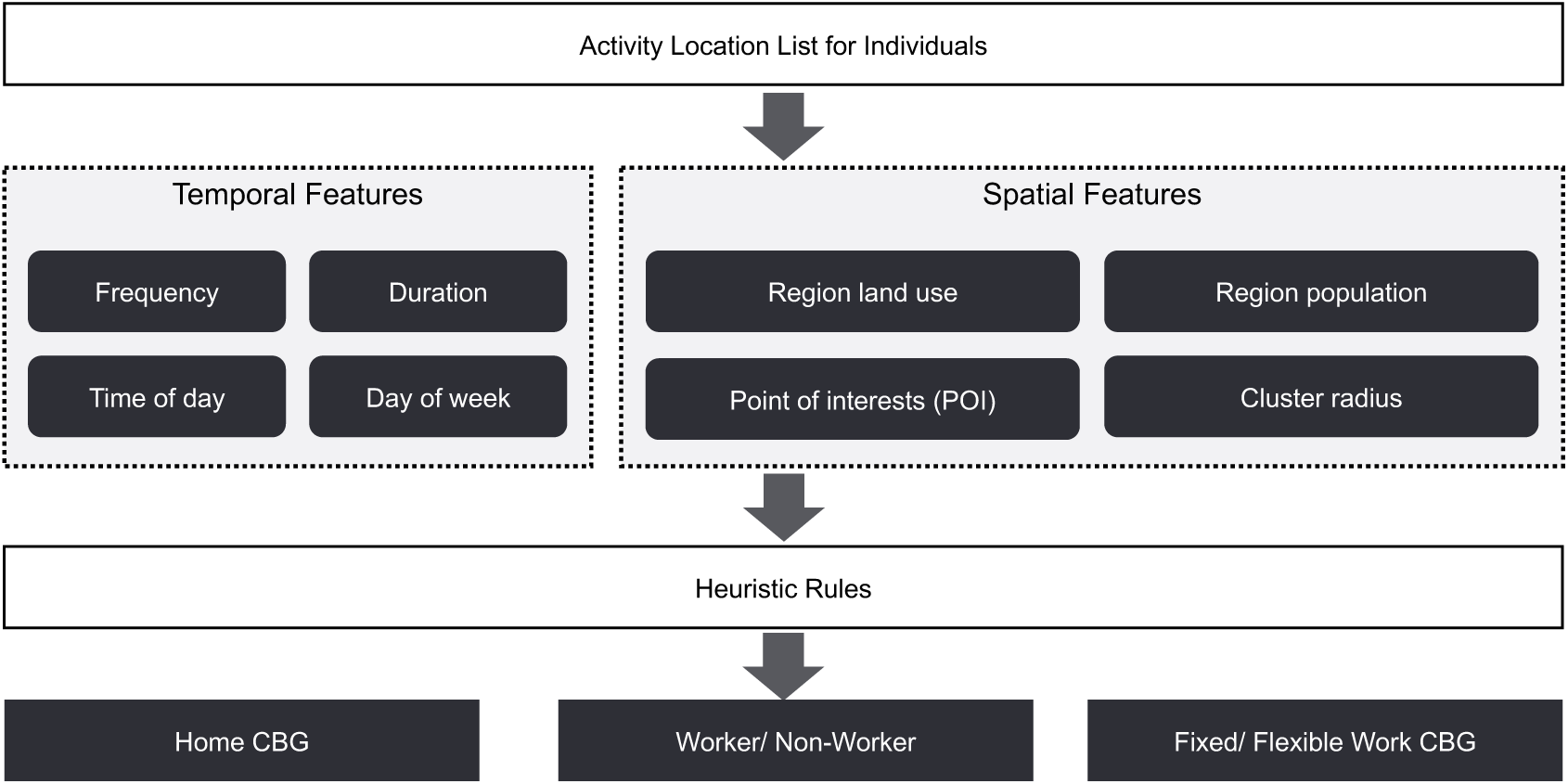
Home/work CBG imputation methodology

**Figure 6.**
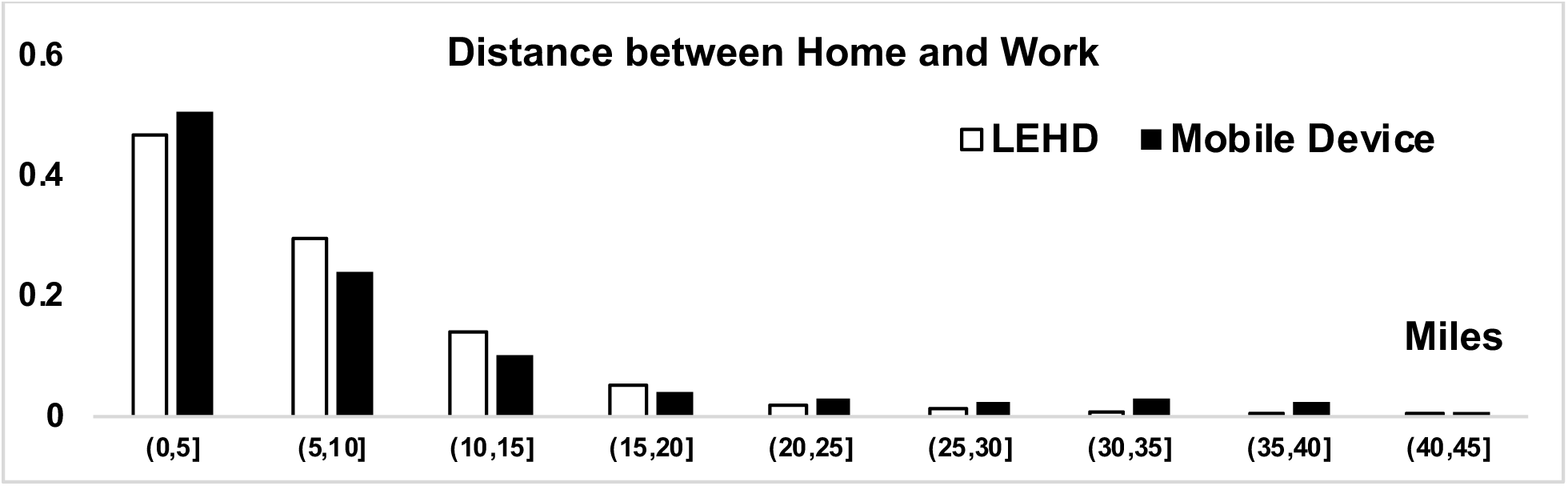
Validation of home and work imputation against LEHD

### 2.3. Imputation

#### 2.3.1. Mode Imputation

Our research team developed a jointly trained single-layer model and deep neural network ^27^ for travel mode detection of this project. This model combines the advantages of both types of models to be able to make sufficient generalizations using a multi-layer DNN and capture the exceptions using the wide single-layer model. The datasets used for training the model were collected from the incenTrip mobile phone app ^28^, developed by the authors, where the ground truth information for car, bus, rail, bike, walk, and air trips was collected. To effectively detect the travel mode for each trip, feature construction is critical in providing useful information. Travel mode-specific knowledge is needed to improve the detection accuracy. In addition to the traditional features used in the literature (e.g. average speed, maximum speed, trip distance, etc.), we also integrated the multi-modal transportation network data to construct innovative features in order to improve the detection accuracy based on network data integration. The wide and deep learning method utilized in this study achieved over 95% prediction accuracy for drive, rail, air, and non-motorized, and over 90% for bus modes on test data. We have applied the trained algorithms on the location dataset to obtain multimodal trip rosters (see **Figure 7** that shows raw location data points by different travel modes). The resulting mode shares show a decent match with the available travel surveys at both national and metropolitan levels.

**Figure 7.**
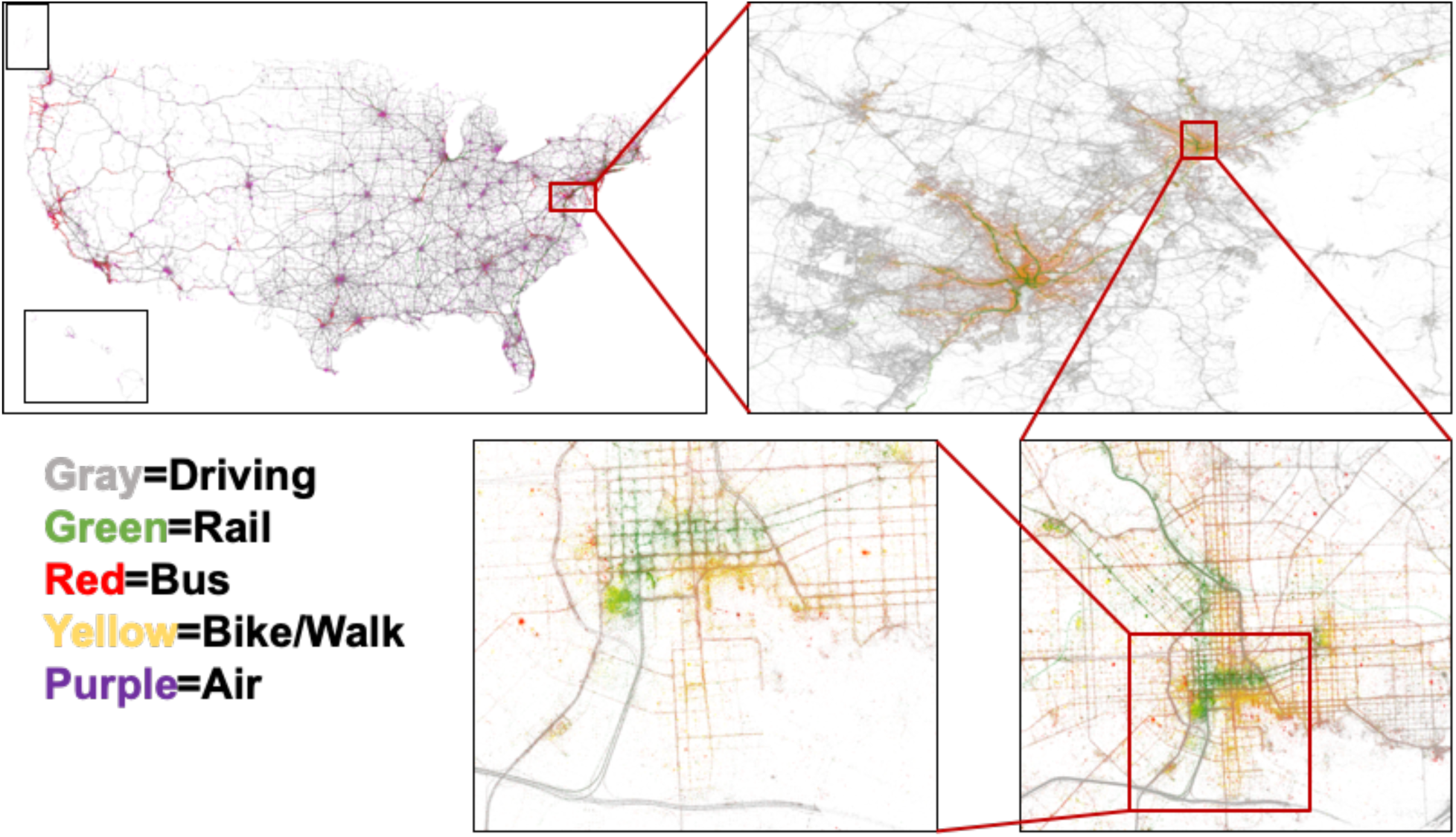
Demonstration of the multi-modal travel patterns

#### 2.3.2. Purpose Imputation

In the Home and work CBG Identification, we described how home and work CBGs can be identified. Other purposes can be directly identified through spatial joint of trip end locations and point of interest (POI) data. We have used a popular commercial POI dataset that includes more than forty million records for the U.S. For each trip end, we first filter all POIs that are located within a 200-meter radius of the trip-end. Next, we identify the trip purpose by the POI type of the closest POI.

#### 2.3.3. Socio-Demographic Imputation

Due to privacy concerns, mobile device location data contain very little ground truth information about the device owners. However, it is essential to understand how representative the sample is and how different segments of the population travel. The state-of-the-practice method is to assign either the census population socio-demographic distribution or the public use microdata sample (PUMS) units to the sample devices within the same geographic area based on the imputed home locations. More advanced socio-demographic imputation methods utilize travel patterns and visited POI types to impute the socio-demographics. These methods require a significant amount of computation, as various features from different databases should be calculated and used. In order to balance the computations and conduct a timely analysis for the pandemic, we have used the state-of-the-practice method and assigned socio-demographic information to the anonymized devices based on the census socio-demographic distribution of their imputed CBG. Five-year American Community Survey (ACS) estimates for 2014 to 2018 from the U.S. Census Bureau can be used to obtain median income, age distribution, gender distribution, and race distribution for each U.S. CBG ^29^. For each device, we used Monte-Carlo simulation ^30^ to draw from the age, gender, and race distribution at the device’s imputed home CBG. We also assigned the CBG’s median income to the device.

### 2.4. Weighting

The sample data needs to be weighted to represent population-level statistics. First, the devices available in our dataset are a sample of all individuals in the population, so we need to apply device-level weights. Second, for an observed device, only a sample of all trips may be recorded, so trip-level weights are also needed. For the sake of timeliness, we have applied simple weighting methods to obtain county-level device weights and state-level trip weights. In order to obtain device-level weights, we have used the home county, obtained from the imputed home CBG information. The weight for each device is equal to the number of devices observed in the device’s imputed home county divided by the population of the county, so all devices residing in a county would have the same device-level weight. For instance, if our sample includes 100 devices in a county with a population of 2,000, each device would be assigned a weight of 20. Population of each county can be obtained from the U.S. Census Bureau. For trip-level weights, we have calculated number of trips per person (trip rate) for each state during an average weekday in the first two weeks of February from our sample. We have also calculated this trip rate number for each state from NHTS2017. We have used a single trip rate for all trips generated from each state, equal to the NHTS trip rate divided by our observed trip rate.

## 3. PLATFORM OVERVIEW

The COVID-19 Impact Analysis Platform, available at data.covid.umd.edu provides data and insights on COVID-19’s impact with daily data updates. The research team are exploring how social distancing and stay-at-home orders are affecting travel behavior, spread of the coronavirus, and local economies. Through this interactive analytics platform, we are making our data and research findings available to other researchers, agencies, non-profits, media, and the general public. The platform will evolve and expand over time as new data and impact metrics are computed and additional visualizations are developed. **Table 1** shows the current metrics available in the platform at the national, state, and county levels in the United States with daily updates. **Figure 8** illustrates the platform.

**Table 1.** List of metrics available on the COVID-19 impact analysis platform

| Current Metrics | Description |
| --- | --- |
| social distancing index | An integer from 0~100 that represents the extent residents and visitors are practicing social distancing. “0” indicates no social distancing is observed in the community, while “100” indicates all residents are staying at home and no visitors are entering the county.<br>It is computed by this equation:<br>$\text{social distancing index} = 0.8 * [\% \text{staying home} + 0.01 * (100 - \% \text{staying home}) * (0.1 * \% \text{reduction of all trips compared to pre-COVID-19 benchmark} + 0.2 * \% \text{reduction of work trips} + 0.4 * \% \text{reduction of non-work trips} + 0.3 * \% \text{reduction of travel distance})] + 0.2 * \% \text{reduction of out-of-county trips}$ |
| % staying home | Percentage of residents staying at home (i.e., no trips more than one mile away from home) |
| #trips/person | Average number of trips taken per person. |
| % out-of-county trips | The percent of all trips taken that travel out of a county. Additional information on the origins and destinations of these trips at the county-to-county level is available, but not currently shown on the platform. |
| miles traveled/person | Average person-miles traveled on all modes (car, train, bus, plane, bike, walk, etc.) |
| #work trips/person | Number of work trips per person (where a “work trip” is defined as going to or coming home from work) |
| #non-work trips/person | Number of non-work trips per person. (e.g. grocery, restaurant, park, etc.). Additional information on trip purpose (restaurant, shops, etc.) is available, but not currently shown on the platform. |
| #COVID-19 cases | Number of new confirmed COVID-19 cases from the Johns Hopkins University’s GitHub repository. |
| population | Number of residents in a nation, state, or county as reported from the national Census database. |

**Figure 8.**
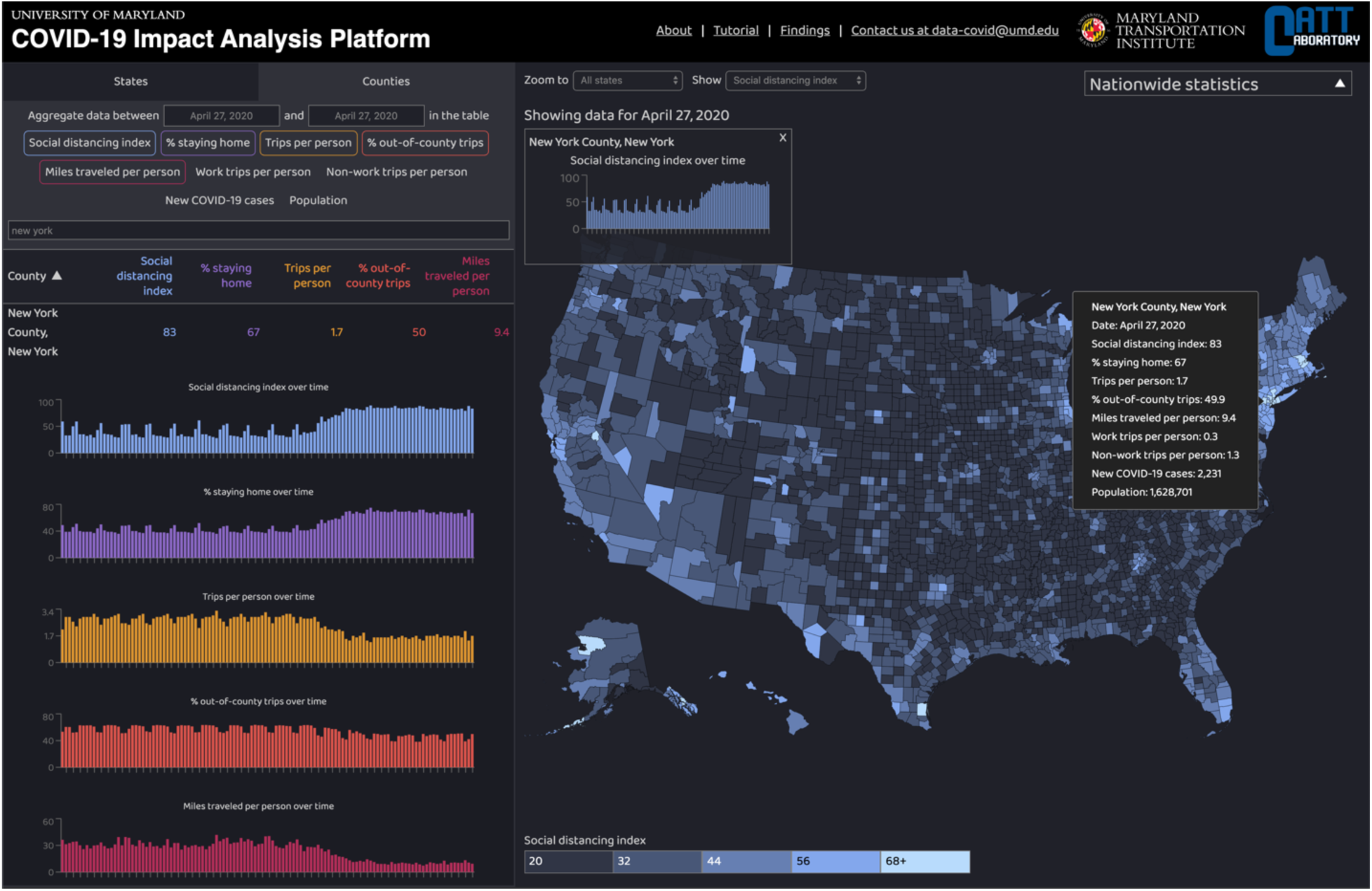
Platform illustration

## 4. CONCLUSION

The Integrated dataset compiled by our research team shows how the nation and different states and counties are impacted by the COVID-19 and how the communities are conforming with the social distancing and stay-at-home orders issued to prevent the spread of the virus. The platform utilizes privacy-protected anonymized mobile device location data integrated with healthcare system data and population data to assign a social distancing score to each state and county based on derived information such as percentage of people who are staying home, average number of trips per person and average distance traveled by each person. As the next steps, the research team is integrating socio-demographic and economic data into the platform to study the multifaceted impact of COVID-19 on our mobility, health, economy, and society.

## Data Availability

All the data are available at data.covid.umd.edu

https://data.covid.umd.edu/

## ACKNOWLEDGMENT

We would like to thank and acknowledge our partners and data sources in this effort: (1) Amazon Web Service and its Senior Solutions Architect, Jianjun Xu, for providing cloud computing and technical support; (2) computational algorithms developed and validated in a previous USDOT Federal Highway Administration’s Exploratory Advanced Research Program project; (3) mobile device location data provider partners; and (4) partial financial support from the U.S. Department of Transportation’s Bureau of Transportation Statistics and the National Science Foundation’s RAPID Program.

